# Comparative Analysis of Detection Efficacy Across Different Nucleic Acid Testing Modalities in Blood Centers

**DOI:** 10.1101/2025.09.05.25334984

**Authors:** Ge Xuelong, Qian Mingming, Yang Xiaohua, Zhang Liwei, Wang Yanbin

**Affiliations:** Hebei Provincial Blood Cente

**Keywords:** ID-NAT, MP-NAT, Testing Efficacy, NAT Yield, Cost-effectiveness

## Abstract

**Background and Objectives:** Blood center Nucleic Acid Testing (NAT) laboratories primarily utilize Individual testing(ID-NAT) and minipool testing(MP-NAT) modes. ID-NAT offers higher sensitivity, while MP-NAT is more cost-effective. While direct comparisons of the two modes exist, the efficacy of a parallel implementation combining both modes has been less explored.

**Materials and Methods:** This laboratory employs both testing systems in parallel. From March to August 2024, the ID-NAT system was suspended for equipment upgrades. NAT indicators during this suspension period were retrospectively analyzed and compared with data from the preceding seven years to assess impacts on economic benefits and positive yield.

**Results:** During the suspension period, 96,941 specimens were tested. The NAT yield rate was 0.0536%, NAT resolution reactive rate was 0.8624%, NAT resolution efficiency rate was 49.06%, MP-NAT reagent utilization rate was 82.35%, NAT invalid rate was 1.5228%, and NAT invalid pool rate was 0.0328%. Comparisons with the same period in 2023 and other months in 2024 showed no statistically significant differences (p > 0.05). Testing costs decreased by approximately 28.57%. When compared to historical data from the same period, the NAT yield rate,and MP-NAT yield rate, showed statistically significant differences (p < 0.05).

**Conclusion:** Differences exist in NAT yield detection between the parallel ID-NAT/MP-NAT mode and the single MP-NAT mode, requiring cautious interpretation. Economically, the parallel mode demonstrated superior performance.

## 1. Introduction

Since the implementation of NAT in domestic blood center laboratories in 2010, two main testing systems have emerged: ID-NAT and MP-NAT. The representative ID-NAT systems, based on TMA technology, perform combined detection of HBV DNA, HCV RNA, and HIV RNA on single specimens, followed by discriminatory testing for reactive samples ^[1]^ . MP-NAT systems use PCR technology to pool multiple specimens (e.g., 6-in-1 or 8-in-1) into one tube for initial testing; reactive pools undergo resolution testing ^[2,3]^ . Theoretically, factors like sample volume result in a lower detection limit for ID-NAT systems compared to MP-NAT, leading to differences in nucleic acid positive detection ^[4]^ . Most laboratories adopt a single testing mode, whereas our laboratory operates a parallel ID-NAT/MP-NAT mode. Specimens are triaged based on ELISA results: HBsAg, HIV Ag/Ab, and anti-HCV positive samples are not subjected to NAT; ELISA-negative samples enter the MP-NAT system; samples with pending or incomplete ELISA results enter the ID-NAT system. The triage strategy is illustrated in Figure 1. This strategy prevents ELISA-positive specimens from causing pool reactivity requiring resolution, reducing the risk of laboratory contamination. From March to August 2024, our laboratory suspended ID-NAT for system upgrades. During this period, all NAT specimens were processed by the MP-NAT system. Due to reporting time constraints, some specimens entered MP-NAT without ELISA results, potentially introducing positive specimens into the MP-NAT system and altering the overall system’s detection limit ^[4,5,6]^ , which may impact NAT efficacy. We retrospectively analyzed NAT results from this period to investigate the impact of the testing mode change on NAT efficacy.

**Figure 1:**
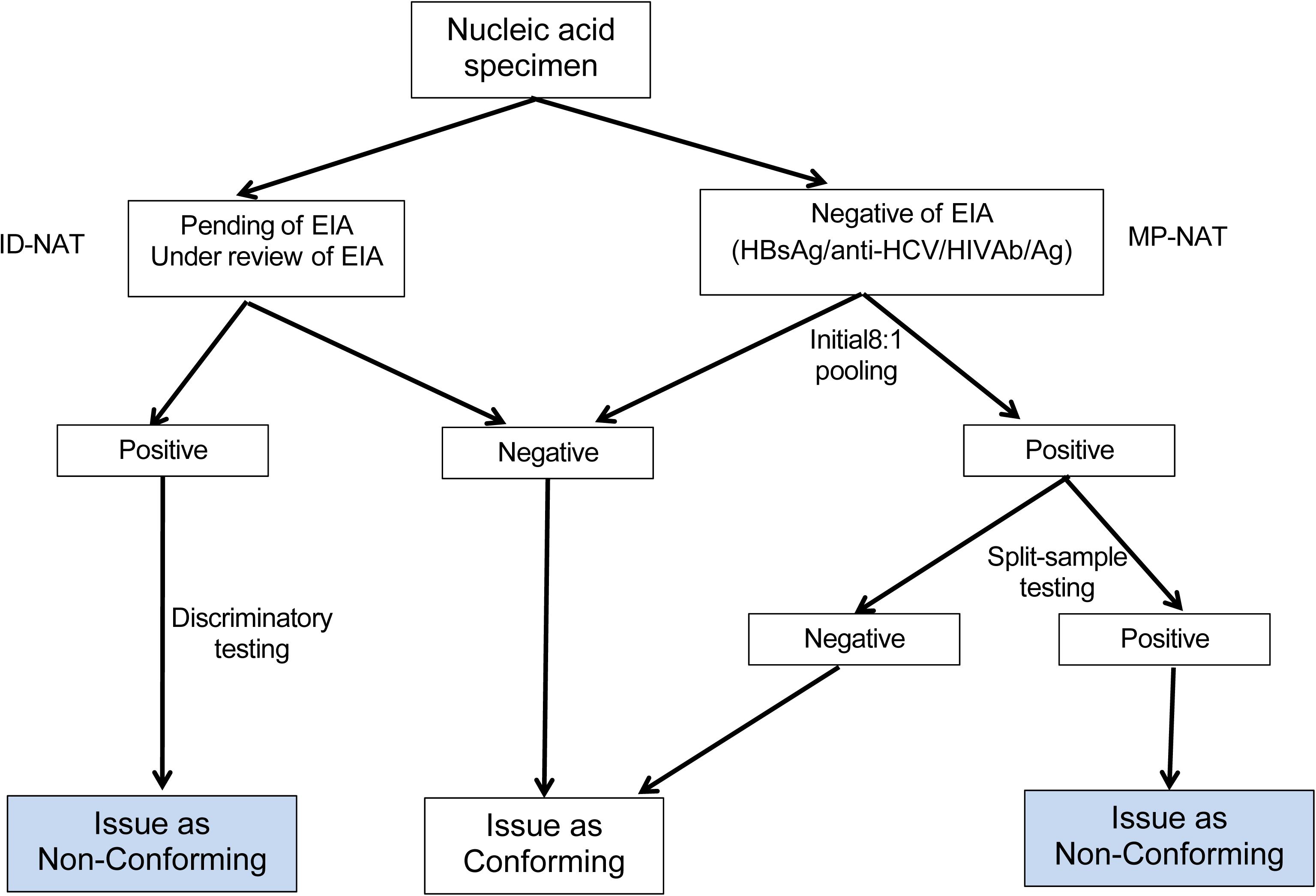

## 2. Materials and Methods

### 2.1 Specimen Source

Blood donor specimens collected in the Shijiazhuang area from January 1, 2018, to December 31, 2024. All donors met the requirements of the “Health Examination Requirements for Blood Donors” (GB18467-2011). A total of 1,458,657 specimens underwent NAT, with 96,941 tested by MP-NAT between March and August 2024.

### 2.2 Reagents and Instruments

ID-NAT system: Grifols Procleix TIGRIS and Procleix PANTHER systems. Reagent lot numbers: 706740, 706154, etc. MP-NAT system: Huayimei NAT system. Reagent lot numbers: MA20231008, MA20220909, etc. All reagents were used within their validity period.

### 2.3 Methods

NAT specimens were processed using a parallel ID-NAT/MP-NAT mode; the triage scheme is shown in Figure 1. To determine if NAT efficacy during the suspension period differed from other periods, key NAT indicators from this period were compared to the same period in 2023 and to other periods in 2024. To mitigate short-term fluctuations and small sample size effects, indicators were also compared to historical data from the same periods.

### 2.4 Statistical Analysis

Data were processed using Excel and SPSS 20.0 software. Comparisons between two groups used the χ² test. Comparisons with historical data used one-sample t-tests (e.g., annual data from 2018–2024). Prediction of the NAT yield rate employed a linear regression model. Spearman’s coefficient was used for correlation calculations. P < 0.05 was considered statistically significant.

## 3. Results

Terminology and Formulas:

ID-NAT Suspension Period: March to August 2024.

Other Periods in 2024: January-February & September-December 2024.

NAT Yield Specimen: Specimen with non-reactive ELISA but reactive NAT result.

NAT Yield Rate = (Total NAT yield specimens / Total NAT tested specimens) × 100%.

MP-NAT Yield Rate = (MP-NAT yield specimens / Total MP-NAT specimens) × 100%.

NAT Resolution Reactive Rate = (Number of reactive NAT pools / Total number of NAT pools) × 100%.

NAT Resolution Efficiency Rate = (Number of pools yielding a reactive result upon resolution / Number of reactive NAT pools) × 100%.

NAT Invalid Run Rate = (Number of invalid NAT runs / Total number of NAT runs) × 100%.

NAT Invalid Pool Rate = (Number of invalid NAT pools / Total number of NAT pools) × 100%.

Reagent Utilization Rate = (Number of reagent doses used / Number of tests performed) × 100%.

Note: Resolution Reactive Rate and Resolution Efficiency Rate exclude ELISA-positive specimens.

### 3.1 Comparison between Suspension Period and Other Periods : See Table 1

### 3.2 NAT Yield Rate and MP-NAT Yield Rate in 2018-2024: See Table 2

### 3.3 Trend Analysis: Historical data for NAT Yield Rate and MP-NAT Yield

Rate, NAT Resolution Efficiency Rate were divided into two groups: Suspension Period (Mar-Aug) and Other Periods (Jan-Feb, Sep-Dec). Trend line charts were generated.

**Table 1.**
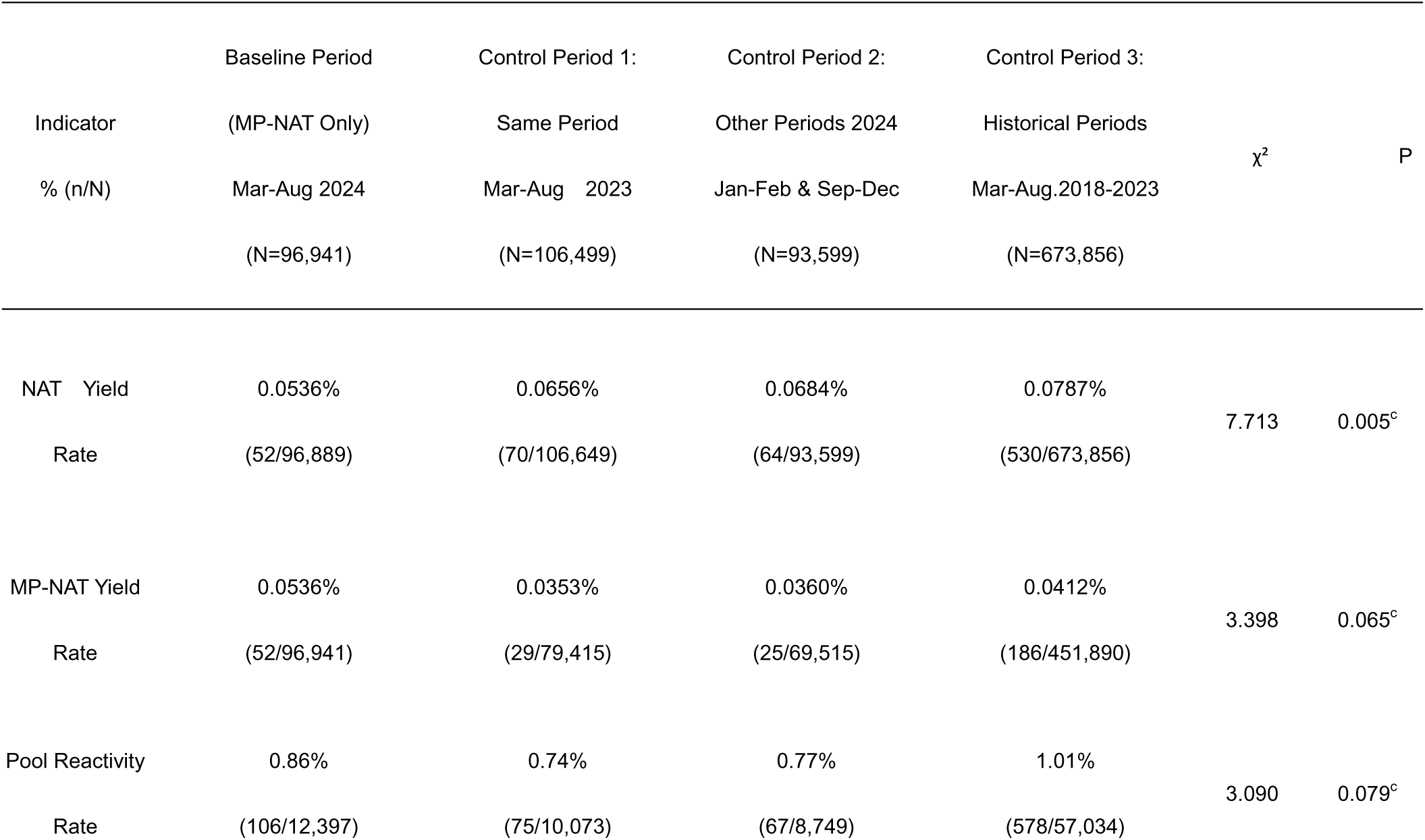
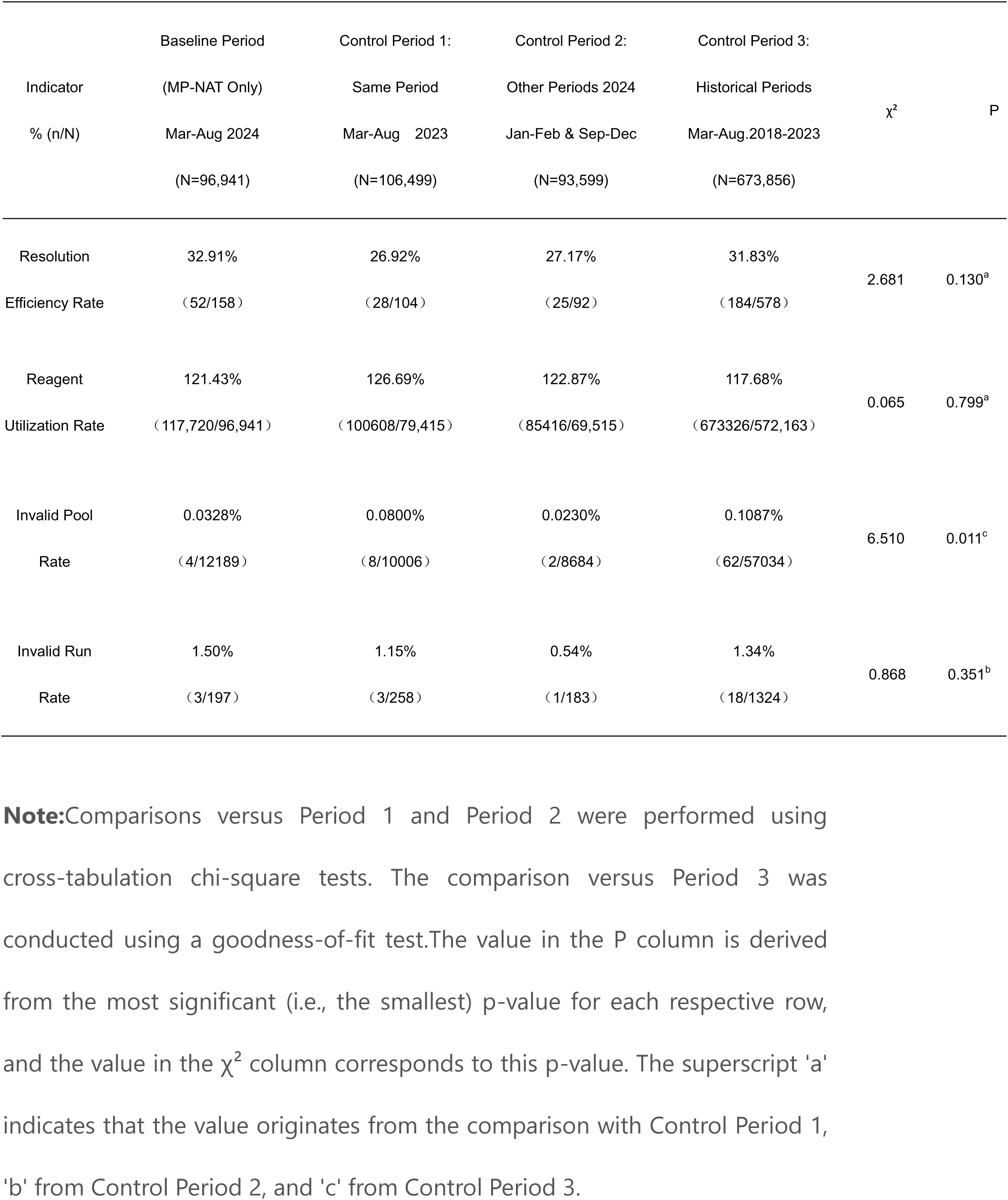
Comparison of Key NAT Efficacy Indicators: Baseline MP-NAT Only Period vs. Control Periods.

**Table 2.**
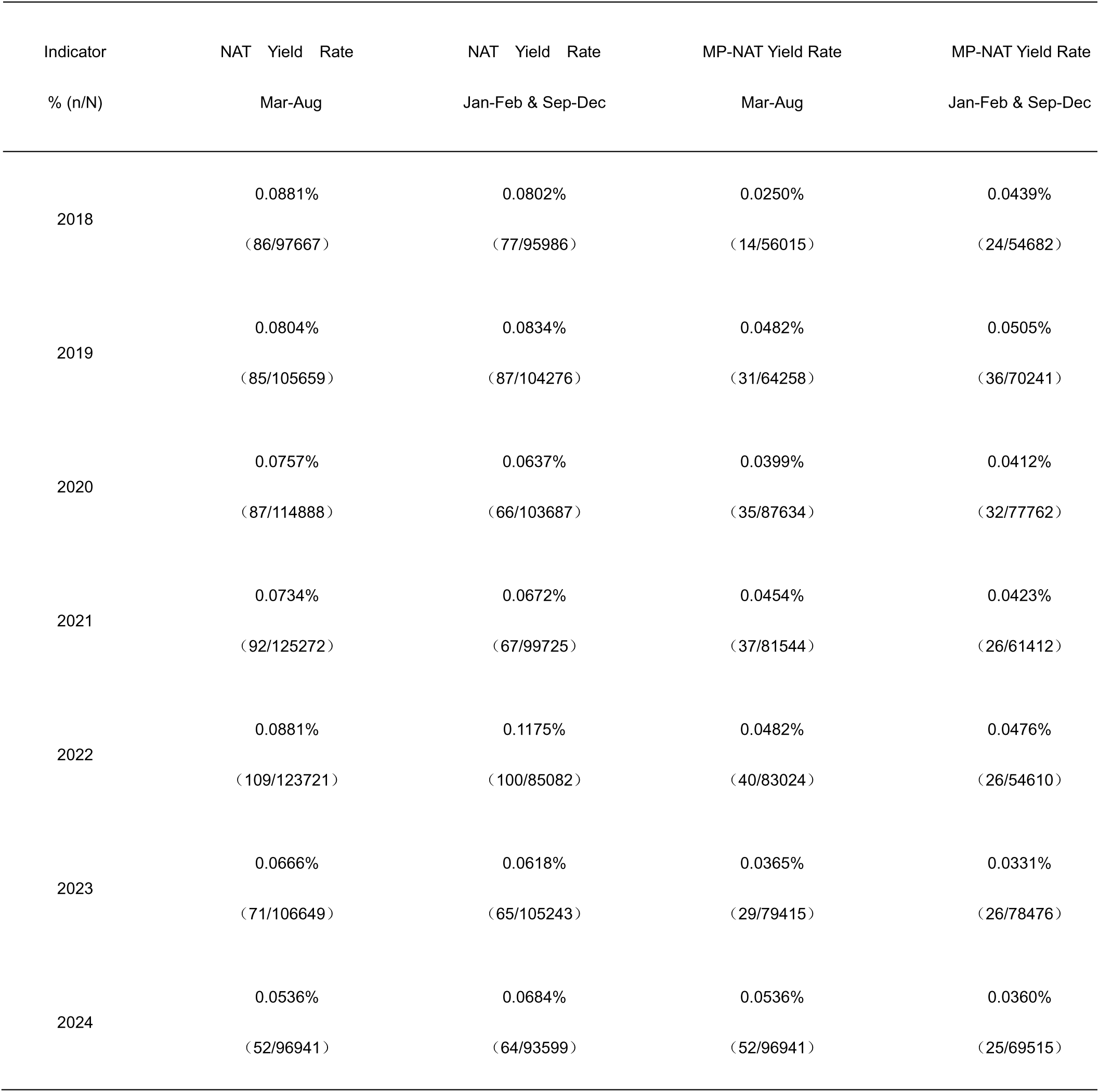
NAT Yield Rate and MP-NAT Yield Rate in 2018-2024.

| Indicator | NAT Yield Rate | NAT Yield Rate | MP-NAT Yield Rate | MP-NAT Yield Rate |
| --- | --- | --- | --- | --- |
| % (n/N) | Mar-Aug | Jan-Feb & Sep-Dec | Mar-Aug | Jan-Feb & Sep-Dec |
| 2018 | 0.0881%<br>( 86/97667 ) | 0.0802%<br>( 77/95986 ) | 0.0250%<br>( 14/56015 ) | 0.0439%<br>( 24/54682 ) |
| 2019 | 0.0804%<br>( 85/105659 ) | 0.0834%<br>( 87/104276 ) | 0.0482%<br>( 31/64258 ) | 0.0505%<br>( 36/70241 ) |
| 2020 | 0.0757%<br>( 87/114888 ) | 0.0637%<br>( 66/103687 ) | 0.0399%<br>( 35/87634 ) | 0.0412%<br>( 32/77762 ) |
| 2021 | 0.0734%<br>( 92/125272 ) | 0.0672%<br>( 67/99725 ) | 0.0454%<br>( 37/81544 ) | 0.0423%<br>( 26/61412 ) |
| 2022 | 0.0881%<br>( 109/123721 ) | 0.1175%<br>( 100/85082 ) | 0.0482%<br>( 40/83024 ) | 0.0476%<br>( 26/54610 ) |
| 2023 | 0.0666%<br>( 71/106649 ) | 0.0618%<br>( 65/105243 ) | 0.0365%<br>( 29/79415 ) | 0.0331%<br>( 26/78476 ) |
| 2024 | 0.0536%<br>( 52/96941 ) | 0.0684%<br>( 64/93599 ) | 0.0536%<br>( 52/96941 ) | 0.0360%<br>( 25/69515 ) |

### 3.6 Reagent Cost-Benefit Calculation

The cost ratio per specimen for MP-NAT versus ID-NAT in our lab is approximately 3:7. Routine testing volume ratio is approximately 7:3 (MP-NAT : ID-NAT). During the suspension period, all specimens underwent MP-NAT.

With no significant difference in reagent utilization, cost savings were calculated as:

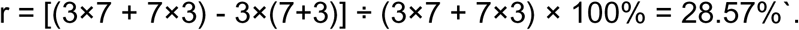

This indicates that the single MP-NAT mode during the suspension period saved approximately 28.57% in reagent costs.

## 4. Discussion

Differences exist between ID-NAT and MP-NAT in the detection limits and timeliness for HIV RNA, HCV RNA, and HBV DNA ^[6,7]^ , with ID-NAT systems generally having lower detection limits than MP-NAT systems ^[8]^ . The 95% detection limits for the ID-NAT system (Procleix) are: HIV RNA-1: 18 (15.0-23.5) IU/ml, HIV RNA-2: 10.4 (8.9-12.6) IU/ml, HCV RNA: 3 (2.5-3.9) IU/ml, HBV DNA: 4.3 (3.8-5.0) IU/ml ^[1,9]^ . These are lower than the Huayimei manufacturer’s claimed limits in resolution mode (42.3 IU/ml, 12.5 IU/ml, 4.2 IU/ml) and even higher in initial MP-NAT mode. Low viral load HBV DNA infection is a major residual risk in blood screening ^[10]^ , making comparisons between single MP-NAT and parallel modes practically significant. Direct comparative studies are limited, as most Chinese blood center NAT labs only use MP-NAT system (e.g., 10 of 12 labs in Hebei Province) ^[11]^ . This contrasts with high-income regions like Australia ^[12]^ and France ^[13]^ , where ID-NAT predominates. Labs running both systems in China triage specimens based on origin or ELISA results, complicating direct efficacy comparisons. Our lab’s suspension of ID-NAT in Mar-Aug 2024, processing all specimens via MP-NAT, provided an ideal model to study the efficacy differences.

Economically, the MP-NAT system’s advantage is clear ^[14,15]^ . For example, Dongguan Blood Center reported ID-NAT equipment runtime and reagent costs were 3.5-5.9 times higher than MP-NAT ^[16]^ . Our lab’s single MP-NAT mode cost during the suspension period was significantly lower than during the parallel period, saving 28.57%. Reagent utilization during suspension increased by 5.2 and 1.4 percentage points compared to the same period last year and other 2024 periods, respectively (not significant). This is attributed to larger batch sizes improving utilization. Slight increases in NAT Resolution Reactive Rate and NAT Invalid Run Rate did not offset the impact of larger batch sizes.The parallel testing mode holds greater reference significance and practical value for blood banking services in low- and middle-income countries compared to the single ID-NAT mode.

During the suspension, time constraints led to some ELISA single-reagent positive or pending-result specimens entering NAT, increasing the likelihood of resolution reactions. Data reflected this: 9 reactive pools were caused by ELISA-positive specimens during suspension vs. 2 in both comparison periods. Monthly environmental monitoring showed zero positive detections in both years, indicating minimal lab environmental impact.

The NAT Resolution Reactive Rate during suspension (0.8699%) was higher than in other 2024 periods (0.7717%) and the same 2023 period (0.7301%). The NAT MP-NAT Yield Rate was also higher. Conversely, the NAT Yield Rate was lower during suspension. Other indicators (Resolution Efficiency Rate, MP-NAT Reagent Utilization, Invalid Pool Rate, Invalid Run Rate) showed no significant differences (P > 0.05, see Tables 1). While differences existed in key indicators between modes, they lacked statistical significance.

During the ID-NAT suspension period, no statistically significant differences were observed in key metrics—including the NAT yield rate, MP-NAT yield rate, NAT resolution reactive rate, NAT resolution efficiency rate, MP-NAT reagent utilization rate, NAT invalid well rate, and NAT invalid run rate—when compared with either the same period in the previous year or other periods within the same year (p > 0.05).

However, when the analysis was expanded to a longer timeframe (spanning the past seven years) as shown in Table 1, statistically significant differences emerged in certain metrics when compared to earlier benchmarks (p < 0.05). A significant difference was observed in the NAT invalid well rate (p = 0.011), attributable to a mechanical failure in the MP-NAT system in July 2020. Due to pandemic-related constraints, repairs were delayed, resulting in 20 instances of low internal control signals that led to invalid determinations. After excluding data from this month, the difference in the NAT invalid well rate was no longer significant.

The most critical metric, the NAT yield rate, demonstrated notable differences. As more clearly illustrated in the trend chart in Figure 2, the NAT yield rate during the 2024 suspension period was 0.0536% (95% CI: 0.0516%– 0.0886%). Although the difference in the MP-NAT yield rate was less pronounced, it also became apparent when data from additional periods were incorporated. A goodness-of-fit test (Table 1) indicated that the difference between the 2024 suspension period and historical data was not statistically significant (p = 0.065).

**Figure 2:**
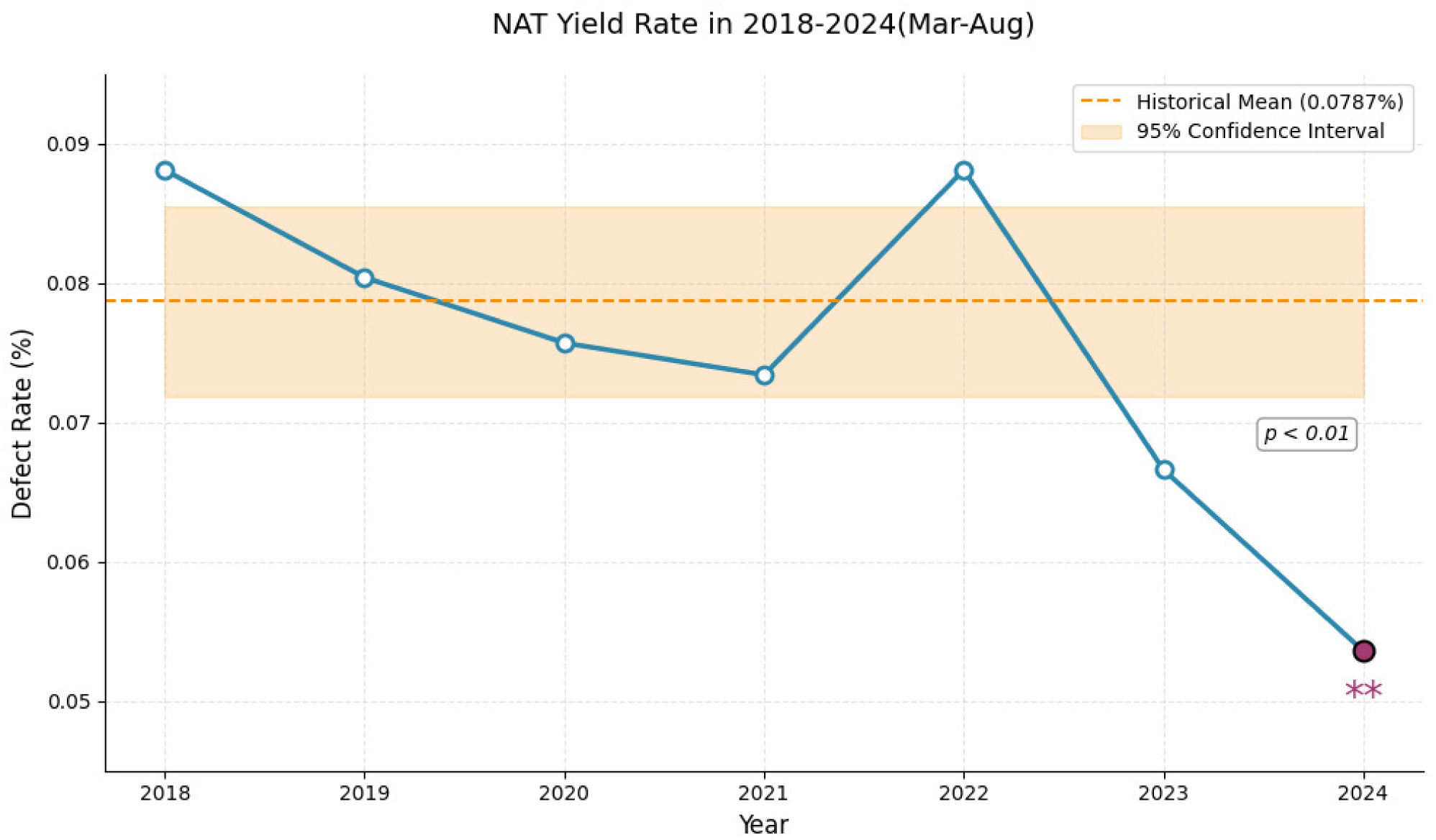
NAT Yield Rate Comparison (2018-2024).

The MP-NAT yield rates from January–February and September– December of each year were grouped together and compared against the rates from March–August in a trend analysis, as shown in Figure 3. After excluding 2018 data, the MP-NAT yield rates for these two period groups were highly correlated from 2019 to 2023 (r = 0.941). However, a clear divergence emerged in 2024. This suggests that relying solely on short-term analyses may underestimate true process variations or lead to misjudgments regarding quality status, underscoring the critical importance of a long-term perspective in quality management and continuous improvement.

**Figure 3:**
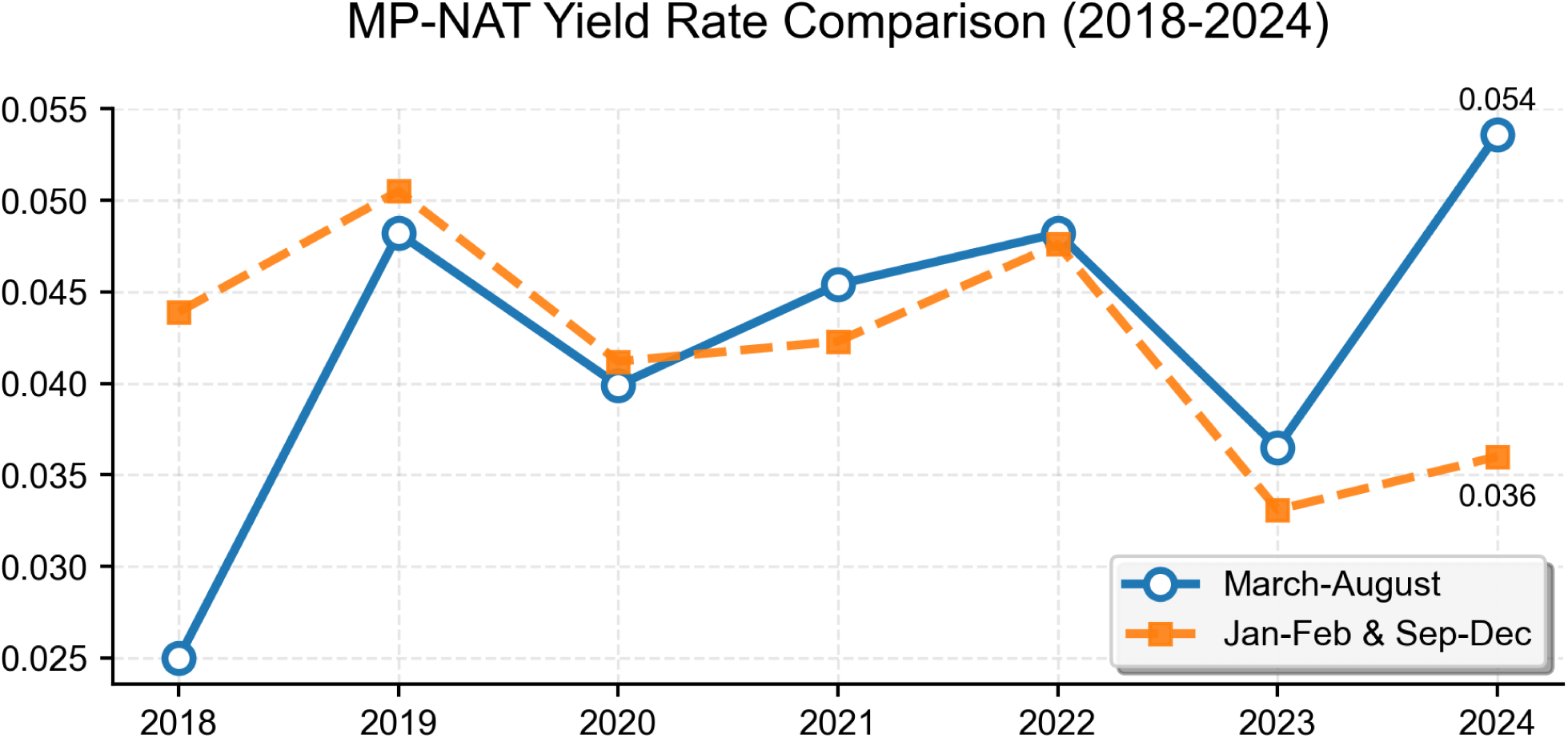
MP-NAT Yield Rate Comparison (2018-2024).

The significant rise in MP-NAT Yield Rate during suspension is likely linked to changes in specimen triage strategy, increased total/batch specimen volume, and inclusion of ELISA-positive specimens^[17,18]^ , requiring further confirmation. Changes in HBV/HCV/HIV prevalence and donor demographics ^[19]^ are also potential factors. Studies indicate higher NAT yield risk among males, aged 45-60, with lower education (farmers in Weinan, Shaanxi) ^[20]^ ; males >35 years in Suzhou for HBV NAT yield ^[21]^ ; similar demographics in Hainan ^[22]^ ; and males in India ^[3]^ and Nigeria ^[10]^ .We propose integrating demographic risk factors (e.g., male donors >45 years with lower education ^[20–22]^) into triage algorithms – directing such specimens to ID-NAT while maintaining low-risk specimens in MP-NAT pools. Dynamic threshold adjustments based on real-time prevalence data, as piloted in Southern China ^[23]^, could further enhance safety without eroding cost savings. ^[23,24,25]^ .

The parallel ID-NAT/MP-NAT mode in blood center NAT labs is less cost-effective than single MP-NAT mode. Differences exist in NAT yield detection, and conclusions vary depending on the comparison timeframe. A key limitation is our inability to fully disentangle the impact of triage strategy changes from inherent mode sensitivity differences. Future multi-center collaborations standardizing triage protocols across parallel and single-mode systems would strengthen causal inference. Additionally, long-term residual risk modeling comparing both modes under controlled triage conditions is warranted.

## Data Availability

All data produced in the present work are contained in the manuscript

## Notes

### Competing Interest Statement

The authors have declared no competing interest.

### Funding Statement

This study was funded by Guided Research Project of the Medical Scientific Research Project Plan of Hebei Provincial Health Commission (No. 20250996)

### Author Declarations

The Ethics Committee of Hebei Provincial Blood Center waived ethical approval for this study as it involved the analysis of pre-existing, anonymized data/samples.

